# Assessment of prescribing in under-five pediatric outpatients in Nigeria: an application of the POPI (Pediatrics: Omission of Prescription and Inappropriate Prescription) tool

**DOI:** 10.1101/2022.03.15.22272436

**Authors:** Ufuoma Shalom Ahwinahwi, Valentine U Odili, Destiny Obiajulu Nwachukwu

## Abstract

**Background:** Ensuring the right drug for the right clinical condition in children under five years of age will dramatically reduce morbidity and mortality rates in developing countries where these values are alarmingly high. This study evaluated prescribing in children under the age of five attending pediatric outpatient clinics at three Central hospitals in Delta State, Nigeria, using the Pediatics: Omission of Prescriptions and Inappropriate Prescription (POPI) tool.

**Methods:** This was a prospective descriptive study of prescriptions made to children from 0 to 59 months who attended the clinics between August and November 2018.Prescriptions were evaluated using the POPI tool, occurrence of potentially inappropriate prescriptions and prescribing omissions were reported as percentages and inappropriate prescription types and prescription omissions were also reported as frequencies. Relationship between inappropriate prescriptions, omissions of prescriptions, and categorical variables of age group and sex, p <.05 were considered significant.

**Results:** A total of 1,327 prescriptions from the three centers were analyzed. There was a preponderance of infants (> 1 month-12 months of age) in the study (43.0%) and a somewhat even gender distribution. Exactly 29.8% of all the prescriptions studied had at least one occurrence of inappropriate prescription. The use of H1 antagonists with sedative or atropine-like effects accounted for the majority of inappropriate prescriptions (49.5%), while the prescription of drinkable amoxicillin or other antibiotics in doses other than mg was the most frequent omission of prescription (97.2%). There was a significant relationship between the occurrence of inappropriate prescription and age group (p> 0.001).

**Conclusion:** The occurrence of inappropriate prescriptions and omissions of prescriptions was high and effectively detected by the POPI tool. Measures should be taken to improve prescribing in order to reduce morbidity and mortality in children below five years.

## Introduction

Medicines use in children as well as in other age groups should be rational ensuring the right drug for the right clinical condition. This should all the more be emphasized in developing countries where the morbidity and mortality rates are quite alarming for children especially those under five years of age. The pediatric population consist of a vulnerable group of individuals who could be exposed to untoward effects of drug because of the peculiarities associated with pharmacokinetic variations which occur due to their poorly developed organs[1] unlicensed medications and off-label drug use. Irrational prescribing can also result in the occurrence of adverse drug reactions which could in some cases lead to hospitalization. There is paucity of studies on rational drug prescribing in the pediatric population; but its importance is gaining increasing recognition [2]. Some tools have been developed to enhance appropriate prescribing in the adult population and their use have been well established. These include the Prescribing Optimally in Middle –aged Peoples’ Treatment (PROMPT), Screening Tool for Older Persons Prescription(STOPP) / Screening Tool to Alert doctors to Right Treatment (START) and Beers Criteria[3-6]. The older population has had a number of these tools and have received attention because they are perceived to be very vulnerable because of the decline in the function of their organs and also the presence of comorbidities resulting in polypharmacy [7]. Until 2011, there was no developed Criteria or a tool for determining inappropriate prescribing specifically in children. Prot-Labathe and colleagues developed, Pediatric Omission of Prescription and Inappropriate Prescription (POPI)[8] and a modified version of the POPI which is POPI (UK) was also developed to suit the UK Clinical Practice[9]. Furthermore, the Potentially Inappropriate Prescribing in children (PIPc) indicators was developed in Ireland[10] and more recently the Key Potentially Inappropriate Drugs (KIDs) List developed by the Pediatric Pharmacy Association of the United States[11]. The KIDs List was specifically developed as a tool to identify medication which have a high tendency to cause adverse drug reactions in children.

A number of studies has been done in Nigeria to detect inappropriate prescribing in the elderly using Beers criteria and STOPP/START criteria [12-14]. However, no study has been reported on the detection of inappropriate prescribing in children using prescribing indicators; our search has also revealed no such study in Africa.

The POPI tool was developed in France but was based on United States, United Kingdom and French guidelines [8]. It comprises I05 criteria with 80 inappropriate prescription (IP) and 25 as omissions of prescriptions (OP) on the list, although 104 was stated by the authors’ report [8]. These criteria were categorized majorly into Diverse Illnesses, Digestive Problems, ENT-Pulmonary Problems, Dermatological Problems and Neuro-psychiatric disorders. Potentially inappropriate prescriptions detect misprescribing and overprescribing while potentially inappropriate omissions detect underprescribing (omissions). The POPI had a consensus validation and has been validated with a retrospective study in a French Hospital and a community pharmacy setting

Potentially inappropriate prescriptions and prescription omissions were detected in a hospital as 2.9% and 2.3% and the community pharmacy as 12.3% and 6.15% respectively [15]. The modified (UK) POPI has also been used in the English healthcare setting [9]. The applicability of the POPI tool outside of French setting has however not been ascertained as discovered from published research. It is worthy of note that in the process of writing up this study, the reports of an international validation of the POPI tool was published [16].

Nigeria is the most populous country in Africa, with over 40% of its population below 16 years of age with the mortality rate of children under age five is 138 per 1000 live births [17]. This population will derive utmost benefit from healthcare if medicines prescribed are appropriate because irrational prescribing can in fact worsen the state of health by undue exposure to adverse drug reactions. Delta State in Nigeria has put in place measures to improve on healthcare by providing free Medicare for children below age five since 2007. This laudable initiative can be even more beneficial when prescribing for this age group is appropriate.

This study aims to assess prescribing in under-fives using the POPI tool and specifically to determine incidence of inappropriate prescriptions and omissions of prescriptions using the POPI tool, as well as to determine the conditions mostly associated with inappropriate prescriptions and omissions in this age group.

## Methods

### Study design

This was a prospective cross-sectional study of prescriptions made out to pediatric patients aged between 0 and 5 years.

### Study setting

This study was conducted in three Central hospitals purposively chosen from the three senatorial districts of the state namely, Central hospital Agbor, Central hospital Warri and Central hospital Ughelli between August 2018 and November 2018.The under-five pharmacy unit takes care of drug dispensing for children below the age of five who receive free medical care at the time of the study. The study was conducted at the pharmacy departments where the prescriptions for pediatrics patients were filled.

### Study population

The population studied were children aged 0-59 months. All prescriptions for pediatric patients aged below five years dispensed at the out-patient pharmacies of the surveyed facilities during the period of data collection were included in the study. At the Central Hospital Warri 462 prescriptions dispensed from 20^th^ August – 20^th^ November 2018. At the Central Hospital Agbor 478 prescriptions were dispensed between the 1^st^ of September 2018 and 30th October 2018 and a total of 387 prescriptions were dispensed at the, Central Hospital Ughelli between September 24^th^ – October 30^th^ 2018.

### Data collection

The prescriptions presented at the pharmacy for filling after consultation with doctors were assessed. Patient information which included age, gender, medical diagnosis, and drug prescribed were collected and additional information such as symptoms if available were extracted from prescription sheets and case files. Weight was not recorded for all patients and was therefore not included as data collected. Data was collected for all cases and recorded using a pro forma developed for data extraction. Prescriptions were assessed on individual basis irrespective of whether it was a repeat consultation or follow-up visit during the study period.

Assessment of prescriptions using the POPI tool: all extracted prescriptions were assessed using the POPI tool. Any omissions in the prescription were recorded on the pro-forma and all inappropriate prescriptions were noted. At the end of the data extraction, all information collected were compared with the information contained on those itemized on the POPI tool. The POPI tool used contained 105 criteria as presented by the authors ^[8]^. Some of these criteria which were either not applicable to this population or for which information where not available on the prescriptions were not analyzed as part of the study, these were

1. DEET: ‘30% (max) before 12 years old” ; “50% (max) after 12 years old” (Omission for mosquitoes)
2. IR3535:”20(Max) before 24 months old; “35% (max)after 24 months old. (Omission for mosquitoes).
3. Mosquito nets and clothes treated with pyrethroids(Omission for mosquitoes)
4. Failure to propose a whooping cough booster vaccine for adults who are likely to become parents in the coming months or years (Omission for Cough)
5. Cyproterone +ethinyl estradiol as a contraceptive to allow isotretinon per os (Inappropriate prescription for Acne vulgaris)
6. Contraception (provided with a logbook or diary) for menstruating girls taking isotretinoin (Omission for Acne vulgaris)

Total number of criteria analyzed for this study were 99 of the 105 criteria stated on the POPI tool (English Version)

### Data analysis

Data retrieved were first analysed with the aid of the POPI tool to detect occurrence of Potentially Inappropriate Medicine and Omission of prescription which were recorded on the data collection form as inappropriate medicines and Omissions accordingly. Age of patients were categorized into age groups and reported in percentages. Occurrence of inappropriate medicines and omission of prescription were categorized and reported as either occurrence or non occurrence. Chi square tests was used to determine relationship between categorical variables (age group and gender) and occurrence of inappropriate prescribing and omission of prescription. Statistical Package for Social Sciences version 23 (IBM Corp Armonk, NY:) was used for analysis with statistical significance established at p< 0.05.

## Results

A total of 1327 prescriptions were analysed in all three centres used for the study. Children within the age range of greater than one month to 12 months (infants) made up a higher percentage of hospital visits during the study (570, 43.0%). And about half the population studied were female (665,50.1%). Table 1 shows details of demographics

**Table 1.** Demographics of study population

| Characteristic |  | Central Hospital,<br>Warri<br>n=462 | Central Hospital,<br>Agbor<br>n= 478 | Central Hospital,<br>Ughelli<br>n=387 | Total<br>N=1327 |
| --- | --- | --- | --- | --- | --- |

|  |  | F (%) | F (%) | F (%) | F (%) |
| --- | --- | --- | --- | --- | --- |
| **Age Group** | 0-1month | 13(2.8) | 27(5.6) | 35(9) | 75(5.7) |
|  | >1 month-12 months | 250 (54.1) | 167(34.9) | 153(39.5) | 570 (43.0) |
|  | >12 months-36 months | 161(34.8) | 203(42.5) | 156(40.3) | 520(39.2) |
|  | >36 months<br>– 59 months | 38 (8.2) | 81(16.9) | 43(11.1) | 162(12.2) |
| **Gender** | Female | 233(50.4) | 230(48.1) | 202(52.2) | 665(50.1) |
|  | Male | 229 (49.6) | 248(51.9) | 185(47.8) | 662(49.9) |

The occurrence of inappropriate prescription in all the prescriptions studied was 29.8%, with Central hospital, Ughelli being the one with the highest prevalence among all centers. Approximately 48% of all the prescriptions studied at this centre had at least one inappropriate prescription. The overall prevalence of prescription omissions was 55.9% and Central Hospital Agbor had up to 71% of prescription omissions. Table 2.

**Table 2.** Occurrence of Inappropriate prescription and Omission of Prescription at the study centres

|  |  | Central<br>Hospital<br>Warri<br>N (%) | Central<br>Hospital<br>Agbor<br>N (%) | Central<br>Hospital<br>Ughelli<br>N (%) | Total<br>N (%) |
| --- | --- | --- | --- | --- | --- |
| Inappropriate prescription |  |  |  |  |  |
|  | Occurrence | 33 (7.1) | 178 (37.2) | 185 (47.8) | 396 (29.8) |
|  | No Occurrence | 429 (92.9) | 300 (62.8) | 202 (52.2) | 931 (70.2) |
| Omissions of prescription |  |  |  |  |  |
|  | Occurrence | 225 (48.7) | 341 (71.3) | 159 (41.1) | 725 (54.6) |
|  | No Occurrence | 237 (51.3) | 137 (28.7) | 228 (58.9) | 602 (45.4) |

Inappropriate prescriptions occurred across the age groups and gender in the different centres as shown in Tables 3 and 4.

**Table 3.** Occurrence of inappropriate prescription across age groups

| Age group | Occurrence of Inappropriate Prescriptions |  |  |  |  |  |
| --- | --- | --- | --- | --- | --- | --- |
|  | Central Hospital | Central Hospital | Central Hospital | Total | X<sup>2</sup> | p- |

|  | Warri<br>F (%) | Agbor<br>F (%) | Ughelli<br>F (%) | F (%) |  | value |
| --- | --- | --- | --- | --- | --- | --- |
| 0-1 month | 3(9.1) | 8 (4.5) | 9(4.9) | 20(5.1) | 19.66 | <0.001 |
| >1month -12<br>months | 16(48.5) | 88(49.4) | 77(41.6) | 181(45.7) |  |  |
| >12 months -<br>36 months | 14(42.4) | 71(39.9) | 84(45.4) | 169(42.7) |  |  |
| >36months- 59<br>months | 0(0) | 11(6.2) | 15(8.1) | 26(6.6) |  |  |
| Total | 33(100) | 178(100) | 185(100) | 396(100) |  |  |

**Table 4.** Occurrence of omission of prescription across age groups

| Age group | Occurrence Omission of prescription |  |  |  |  |  |
| --- | --- | --- | --- | --- | --- | --- |
|  | Central<br>Hospital | Central<br>Hospital | Central<br>Hospital | Total | X<sup>2</sup> | p- |

|  | Warri<br>F (%) | Agbor<br>F (%) | Ughelli<br>F (%) | F (%) |  | value |
| --- | --- | --- | --- | --- | --- | --- |
| 0-1 month | 5(2.2) | 21(6.16) | 16(10.1) | 42(5.8) | 2.96 | 0.398 |
| >1month -12<br>months | 118(52.9) | 125(36.6) | 58(36.5) | 301(41.7) |  |  |
| >12 months -<br>36 months | 82(36.1) | 147(43.1) | 68(42.8) | 297(40.9) |  |  |
| >36months-59 months | 20(8.8) | 48(14.1) | 17(10.7) | 85(11.7) |  |  |
| Total | 225(100) | 341(100) | 159(100) | 725(100) |  |  |

The frequency of occurrence of inappropriate prescription was statistically significantly higher in the >1 to 12 months age group (181;45.7%), followed by the >12 to 36 months age group (169;42.7%) (p< 001) as shown in table 3. Similarly, prescription omissions were more in the >1 to 12 months age group (301;41.7 %), followed by the >12-36 months age group (297;40.9%), however, the difference was not statistically significant. (p=0.398) as shown in table 4. There was no statistically significant difference in inappropriate prescription (p=0.905) and omission of prescription (p=0.07) with regard to gender in the total population studied. Tables 5 and 6 respectively.

**Table 5.** Occurrence of inappropriate prescribing across gender

| Gender | Occurrence of Inappropriate prescriptions |  |  |  |  |  |
| --- | --- | --- | --- | --- | --- | --- |
|  | Central<br>Hospital Warri<br>F(%) | Central<br>Hospital Agbor<br>F(%) | Central<br>Hospital<br>Ughelli | Total<br>F(%) | X<sup>2</sup> | p-value |

|  |  |  | F(%) |  |  |  |
| --- | --- | --- | --- | --- | --- | --- |
| Female | 18(54.6) | 80(44.9) | 96(51.9) | 194(49.0) | 0.025 | 0.905 |
| Male | 15(45.4) | 98(45.1) | 89(49.1) | 202(51.0) |  |  |
| Total | 33(100) | 178(100) | 185(100) | 396(100) |  |  |

**Table.6.** Occurrence of Omission of Prescription across gender

| Gender | Occurrence of Omission of Prescription |  |  |  |  |  |
| --- | --- | --- | --- | --- | --- | --- |
|  | Central<br>Hospital<br>Warri<br>F(%) | Central<br>Hospital<br>Agbor<br>F(%) | Central<br>Hospital<br>Ughelli<br>F(%) | Total<br>F(%) | X<sup>2</sup> | p-value |
| Female | 103(45.8) | 165(48.4) | 82(51.6) | 352(48.6) | 3.311 | 0.07 |
| Male | 122(54.2) | 176(51.6) | 77(48.4) | 347(51.4) |  |  |
| Total | 225(100) | 341(100) | 159(100) | 725(100) |  |  |

Overall, a total of 459 potentially inappropriate medicines were detected in the 396 prescriptions encountered. Based on the systems in which they occurred, 302 of the 459 (65.8%) inappropriate prescriptions were detected in prescriptions written for ENT/Pulmonary (respiratory) systems. Frequent use of H1 antagonist for upper respiratory tract infections or cough associated with other conditions in children less than 30 months old accounted for 49.5% (227/459) of all inappropriate prescriptions, with 133 of the total 227(58.6%) cases occurring at the Agbor centre. Fifty percent(231) of inappropriate prescribing cases occurred in the Ughelli centre. Prescription of a medication other than paracetamol as a first-line treatment for pain was predominant at the ughelli centre where 68 of 82 (82.9%)of such cases occurred. Details of cases and types of inappropriate prescribing are shown in Table 7.

**Table 7.**
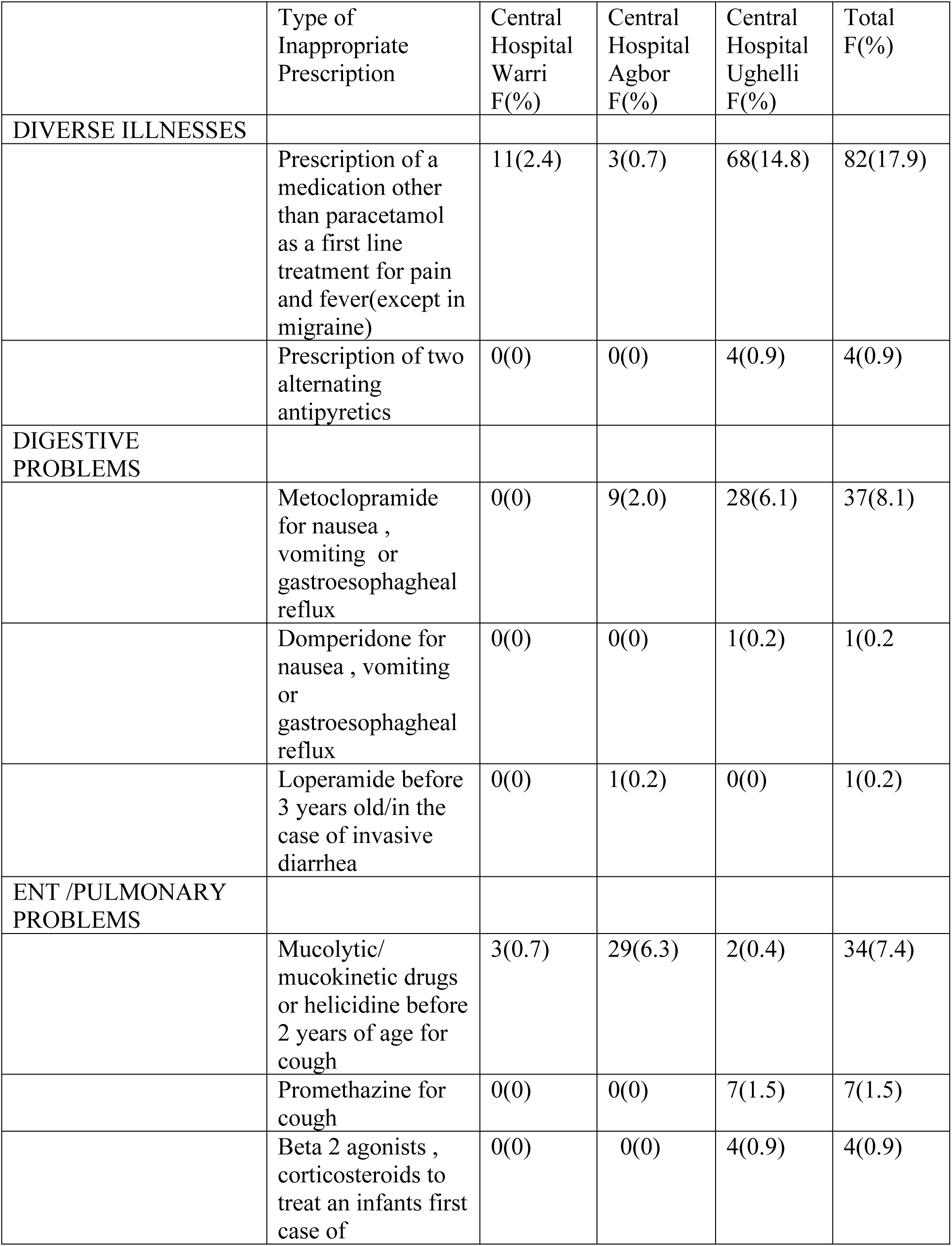
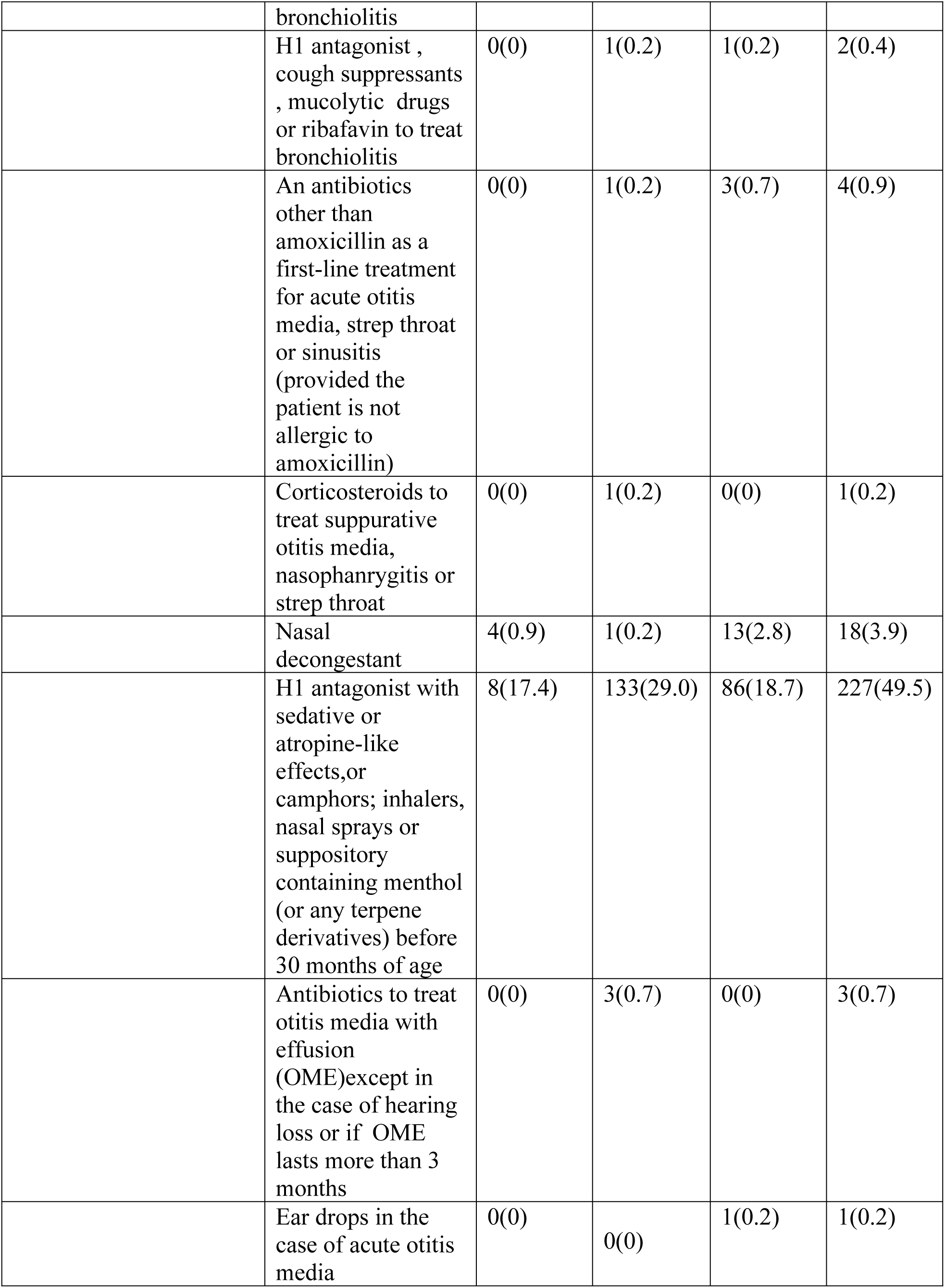
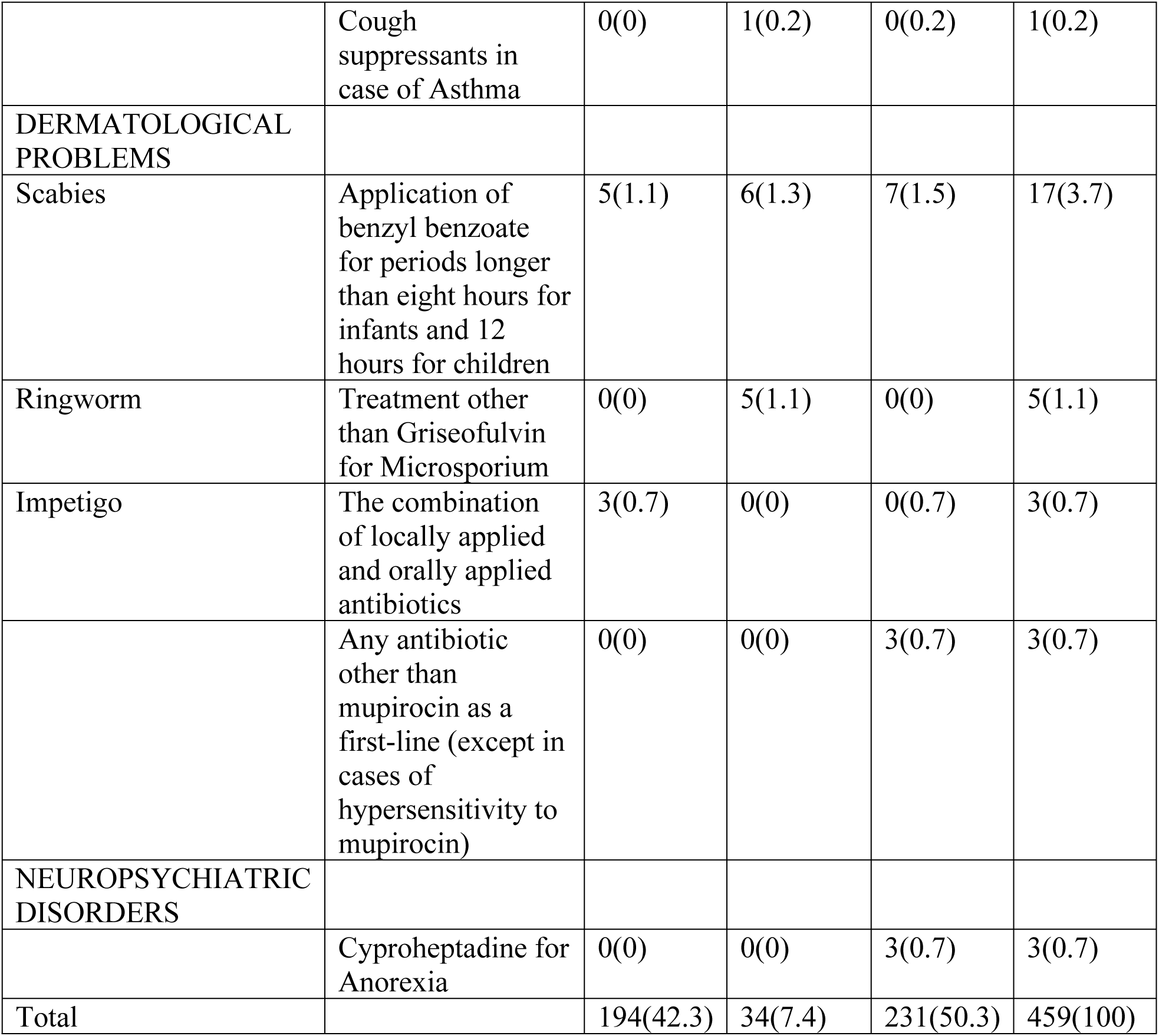
Types of Inappropriate Prescription.

|  | Type of Inappropriate Prescription | Central Hospital Warri F(%) | Central Hospital Agbor F(%) | Central Hospital Ughelli F(%) | Total F(%) |
| --- | --- | --- | --- | --- | --- |
| **DIVERSE ILLNESSES** |  |  |  |  |  |
|  | Prescription of a medication other than paracetamol as a first line treatment for pain and fever(except in migraine) | 11(2.4) | 3(0.7) | 68(14.8) | 82(17.9) |
|  | Prescription of two alternating antipyretics | 0(0) | 0(0) | 4(0.9) | 4(0.9) |
| **DIGESTIVE PROBLEMS** |  |  |  |  |  |
|  | Metoclopramide for nausea , vomiting or gastroesophagheal reflux | 0(0) | 9(2.0) | 28(6.1) | 37(8.1) |
|  | Domperidone for nausea , vomiting or gastroesophagheal reflux | 0(0) | 0(0) | 1(0.2) | 1(0.2) |
|  | Loperamide before 3 years old/in the case of invasive diarrhea | 0(0) | 1(0.2) | 0(0) | 1(0.2) |
| **ENT /PULMONARY PROBLEMS** |  |  |  |  |  |
|  | Mucolytic/ mucokinetic drugs or helcidine before 2 years of age for cough | 3(0.7) | 29(6.3) | 2(0.4) | 34(7.4) |
|  | Promethazine for cough | 0(0) | 0(0) | 7(1.5) | 7(1.5) |
|  | Beta 2 agonists , corticosteroids to treat an infants first case of bronchiolitis | 0(0) | 0(0) | 4(0.9) | 4(0.9) |
|  | H1 antagonist ,<br>cough suppressants<br>, mucolytic drugs<br>or ribavirin to treat<br>bronchiolitis | 0(0) | 1(0.2) | 1(0.2) | 2(0.4) |
|  | An antibiotics<br>other than<br>amoxicillin as a<br>first-line treatment<br>for acute otitis<br>media, strep throat<br>or sinusitis<br>(provided the<br>patient is not<br>allergic to<br>amoxicillin) | 0(0) | 1(0.2) | 3(0.7) | 4(0.9) |
|  | Corticosteroids to<br>treat suppurative<br>otitis media,<br>nasopharyngitis or<br>strep throat | 0(0) | 1(0.2) | 0(0) | 1(0.2) |
|  | Nasal<br>decongestant | 4(0.9) | 1(0.2) | 13(2.8) | 18(3.9) |
|  | H1 antagonist with<br>sedative or<br>atropine-like<br>effects,or<br>camphors; inhalers,<br>nasal sprays or<br>suppository<br>containing menthol<br>(or any terpene<br>derivatives) before<br>30 months of age | 8(17.4) | 133(29.0) | 86(18.7) | 227(49.5) |
|  | Antibiotics to treat<br>otitis media with<br>effusion<br>(OME)except in<br>the case of hearing<br>loss or if OME<br>lasts more than 3<br>months | 0(0) | 3(0.7) | 0(0) | 3(0.7) |
|  | Ear drops in the<br>case of acute otitis<br>media | 0(0) | 0(0) | 1(0.2) | 1(0.2) |
|  | Cough suppressants in case of Asthma | 0(0) | 1(0.2) | 0(0.2) | 1(0.2) |
| **DERMATOLOGICAL PROBLEMS** |  |  |  |  |  |
| Scabies | Application of benzyl benzoate for periods longer than eight hours for infants and 12 hours for children | 5(1.1) | 6(1.3) | 7(1.5) | 17(3.7) |
| Ringworm | Treatment other than Griseofulvin for Microsporum | 0(0) | 5(1.1) | 0(0) | 5(1.1) |
| Impetigo | The combination of locally applied and orally applied antibiotics | 3(0.7) | 0(0) | 0(0.7) | 3(0.7) |
|  | Any antibiotic other than mupirocin as a first-line (except in cases of hypersensitivity to mupirocin) | 0(0) | 0(0) | 3(0.7) | 3(0.7) |
| **NEUROPSYCHIATRIC DISORDERS** |  |  |  |  |  |
|  | Cyproheptadine for Anorexia | 0(0) | 0(0) | 3(0.7) | 3(0.7) |
| **Total** |  | **194(42.3)** | **34(7.4)** | **231(50.3)** | **459(100)** |

**Table 8.** Types of omission of Prescription

|  | Type of Omission of Prescription | Central Hospital, Warri f(%) | Central hospital, Agbor f(%) | Central Hospital, Ughelli, f(%) | Total F(%) |
| --- | --- | --- | --- | --- | --- |
| **DIVERSE** |  |  |  |  |  |

| ILLNESSES |  |  |  |  |  |
| --- | --- | --- | --- | --- | --- |
|  | Asthma inhaler for children | 0(0) | 2(0.3) | 0(0) | 2(0.3) |
|  | Oral rehydration solution during diarrhea and vomiting | 0(0) | 6(0.8) | 0(0) | 6(0.8) |
|  | Doses in mg for drinkable solution of antibiotics in varying concentration | 228(30.9) | 333(45.2) | 155(21.0) | 716 (97.2) |
|  | Decontamination of households linen and clothes and treatment of other family members in cases of scabies | 0(0) | 4(0.5) | 5(0.7) | 9(1.2) |
|  | Tropical treatment combined with oral treatment for microsporium | 1(0.1) | 2(0.3) | 1(0.1) | 4(0.5) |
| Total |  | 229(31.1) | 347(47.1) | 161(21.9) | 737(100) |

A total of 737 omissions of prescriptions occurred in this study, with prescriptions of drinkable amoxicillin and other antibiotics in doses other than in milligrams per ml accounting 716(97.2%) of total omission, this omission of prescription ocured more in Central hospital, agbor with 45.2% of all prescriptions encountered at the centered affected. Only 6 (0.8%) of all omissions involved the prescription of oral rehydration salts in case of vomiting and diarrhea, all of which wnere encountered at Central hospital, Agbor.

## Discussion

This study assessed inappropriate prescription and omissions of prescription in ambulatory pediatric population in Delta state. The particularly high mortality of this age group in Nigeria and vulnerability of under-fives made this of much interest. Our study revealed a high prevalence of potentially inappropriate medicines (PIMs) and potentially prescribing omissions (PPOs) in the pediatric population studied as detected by the POPI tool.

Over a quarter of the prescriptions in our study had at least one inappropriate medicine and more than half had at least an omission of prescription. This is particularly higher than detected in a French Hospital where 2.9% had an inappropriate prescription and 2.7% had an omission of prescription ^[15]^. A similar study done in the UK with a modified POPI tool had 32 out of a total of 400 prescriptions having either inappropriate prescriptions or omission of prescription ^[9]^. However, a high prevalence of inappropriate prescription of 26.4% was detected in a French community pharmacy setting which stock medicines like cough suppressants, domperidone, Rhinotrophyl, which are usually not available in their hospital setting ^[15]^; this finding is comparable to the values obtained from the hospital setting in this study. Similarly, some items such as menthol crystals for inhalation and nasal decongestants were unavailable in the hospitals studied and patients’ caregivers were asked to get these from nearby community pharmacy outlets. Using the PIPc indicators, an overall prevalence of PIPc of 3.5% (95% CI 3.5 to 3.6%) and 11.5% (95% CI 11.4% to 11.7%) by commission and omission respectively among children under 16 years of age across Ireland was detected ^[18]^. Varying prevalence of inappropriate prescribing have been reported in other age groups. The overall prevalence of PIP among middle –aged persons using the PROMPT (Prescribing Optimally in Middle-aged People’s Treatments) criteria was 42.9% ^[3]^ and 21.1%(95% CI 21.0-21.2) in the Republic of Ireland and Northern Ireland respectively. The prevalence of PIP in the Republic of Ireland was much higher than that detected by POPI in the current study while that in Northern Ireland was somewhat lower. A higher prevalence of PIPs was detected in our study when compared with other previous studies among the elderly detected by the STOPP/START criteria where a total of 12.9% and 7% of omission was found among the elderly in an Indian population^[19]^. A comparative study of inappropriate prescribing in the elderly using the 2015 American Geriatric Society updated Beers criteria revealed 35.2% and 29.6% in Nigerian and South African participants respectively (Saka et al,2019). Studies conducted in Nigeria among the elderly has shown a range of PIPs to be 15.7-55.1% using the Beers and STOPP Criteria ^[20] [12] [13] [14]^ and Potential Prescribing Omissions were detected in 4.4% of patient prescriptions ^[21]^. Inappropriate prescribing in children detected with the aid of prescribing tools in Nigeria has not been reported as revealed from search of literature.

Other tools for detecting irrational prescribing in the adult population has been used in more frequent studies with geriatric patients being more in focus. This tools are however not similar as the prevalent disease conditions in both populations are different. Whereas the elderly has cardiovascular and nervous system conditions being more prevalent ^[4]^ children present more with respiratory and gastrointestinal conditions.

Frequent use of H1 antagonists for upper respiratory tract infections or cough associated with other conditions in children less than 30months old accounted for the most encountered inappropriate prescriptions while prescription of amoxicillin or other antibiotics in doses other than in milligrams accounted for the most encountered omission of prescription. The use of H1 antagonist is generally discouraged in young children because of the tendency to cause sedation, dizziness and incoordination in overdose. A New Zealander publication states that sedating antihistamines are contraindicated for all indications in children less than 2 years of age and for cough and colds in children less than 6years of age. (New Zealand Medicines and Medical Devices Authority, 2013). Also, there has not been any evidence found to support the use of sedating antihistamines in treating the symptoms of common colds in children ^[22]^

Mucolytic and mucokinetic cough syrups or helicidine in children below the age of 2 years were frequently prescribed especially at Central Hospital, Agbor,

It was commonly observed in this study that oral amoxicillin and other antibiotics were prescribed in doses other than in mg as this was also the major omission in the French study. The need to specify doses in milligrams cannot be overemphasized as most of these antibiotics are formulated in more than one concentration when reconstituted.

Metoclopramide was commonly used in one particular study centre to control vomiting in the study, it is was however not recommended by the POPI tool due to its tendency to cause extrapyramidal side effects such as tardive dyskinesia and sedation although studies have shown that these are reversible and do not have long term effects ^[23]^.

The prescription of a medication other than paracetamol for pain and fever was mostly encountered at Central Hospital Ughelli with ibuprofen being frequently prescribed, a few occurrences of this inappropriate prescription was also observed at the other study centres.

Although malaria, a common disease condition in children of this age group particularly in the tropics was not included on the POPI tool, control for mosquito which is the vector causing malaria was however among the listed criteria, this item was however not included in the implementation of the use of the POPI because methods for eradication of mosquitoes are a general public health concern and concerted efforts such as the provision of insecticide treated nets are already provided for by the government and DEET and IR3535 are not available for prescribing in our hospitals. Hence assessing the prescriptions for these criteria was needless as they are not always documented on the prescription sheets.

The “prescription of an antibiotic other than amoxicillin as a first-line for acute otitis media, strep throat or sinusitis (provided the patient is not allergic to amoxicillin)” is a criterion which was not effectively assessed as most of the respiratory conditions were not described. Antibiotics were frequently prescribed in the prescriptions studied with amoxicillin occurring as first-line in only 179 of the total of 746 prescriptions on respiratory infections for which antibiotics were prescribed.

The prescription of Oral Rehydration Salts (ORS) for children with symptoms of nausea and vomiting and also for diarrhea is highly commendable in this population. This contrasts the findings in the French study where this was a major PPO. In our study, many prescriptions with malaria and gastroenteritis also had oral rehydration salts prescribed with only a few cases of omissions.

The PIMs were more prevalent in diseases of the ENT/ Pulmonary problems categories with respiratory tract infections commonly diagnosed either alone or comorbid with other conditions giving a total of 304 out of the total 458 (66.4%) PIMs detected in the study. This high occurrence of PIMs is somewhat comparable to the pioneer study conducted in a French hospital setting where 472 of 541 (87.2%) PIMs detected were from prescriptions of ENT/Pulmonary disorders. This is more likely to occur due to the high prevalence of such conditions in the pediatric population which was also evidenced in the French hospital study where 45.2% of all prescriptions had ENT/Pulmonary disorders as pathology. This of course is not comparable in older populations because of the difference in the variations in the pathologies commonly associated in these groups. Notably, many of the prescriptions with malaria as diagnosis also had a high frequency of symptoms of cough and other respiratory symptoms.

The use of antibiotics was common in the prescriptions studied, and in a number of these prescriptions the rationale for their use could not be ascertained. Aside the common occurrence in prescriptions for respiratory tract infections, antibiotics were also commonly prescribed in prescriptions having malaria as the only pathology and in other gastrointestinal /digestive conditions. In majority of these cases, prescription of antibiotics were in doses other than mg /ml which could be misleading as most of these antibiotics are available in more than one concentration.

The use of nasal decongestants was also observed in all the centres with Central Hospital,Ughelli having a higher frequency of occurrence, oral decongestants were not prescribed as they are often in combination with mucolytic/ mucokinetic cough mixtures.

An instance of the use of domperidone prescribed for dyspepsia also occurred in the study, this is a PIM included in the POPI tool. It is associated with Q-T interval prolongation.

A few instances were also observed where promethazine was prescribed for respiratory tract infections either occurring alone or with malaria.

The application of benzyl benzoate for periods longer than eight hours for infants in the treatment of scabies was also detected in our study. This criterion though not included in a newer version of POPI used in the French retrospective study is very much relevant in our study setting.

There was a significant relationship between the occurrence of PIM and age group however this was not so between the occurrence of PPO and age group. Prescription of inappropriate medicines were more in children between 1 month and 12 months than in those greater than 1 year and up to 3 years old. This could have resulted from the observation that these age groups present more at the hospitals compared with the neonates (Birth – 1 month) and young children (> 3years - 5 years). Although, the French retrospective study covered a wider age of children (0-18 years), it reported that children between 0-12 years had a higher risk of presenting with a PIM and no PIM was detected in neonates ^[15]^. Also gender was neither significantly associated with PIM and PPO as both gender were evenly distributed in the two cases.

## Conclusion

The POPI tool was effective in detecting inappropriate prescribing in an under-five ambulatory pediatric population in Nigeria. High incidence of potentially inappropriate medicines and potentially prescription omissions were detected. Inappropriate prescriptions occurred more with prescriptions for ENT/Pulmonary conditions while doses in milligram for drinkable solutions of amoxicillin and other antibiotics was the most frequent occurring omissions of prescription. There was a significant relationship between the occurrence of PIM and age group.

## Data Availability

All relevant data are found within the manuscript and its supporting information files

## Supporting information

S1 Table: Dataset for Agbor Study Centre

S2 Table: Dataset for Warri study centre

S3 Table: Dataset for Ughelli Study centre

